# Mapping Stakeholder Engagement in Endometriosis Care Innovation: Insights from the VendoR Project

**DOI:** 10.64898/2026.04.01.26349826

**Authors:** Somayeh Mahdikhani, Saoirse Cummins, Frances Cleary

## Abstract

**Objectives:** Endometriosis affects approximately 10% of reproductive-age women worldwide, yet care pathways remain fragmented and treatments have limitations. This study aimed to identify and categorize key stakeholders in endometriosis care in Ireland, assess their influence and interest in the digital health initiative, and identify drivers and barriers affecting uptake of innovative approaches to care.

**Methods:** A virtual stakeholder mapping workshop was conducted with participants from healthcare, policy, education, technology, academia, and patient communities. Using a structured MS Teams Whiteboard, participants generated a stakeholder list, positioned stakeholders on an Influence–Interest Matrix, and provided qualitative insights on factors enabling or constraining engagement with digital health innovation.

**Results:** Stakeholders were distributed across all four quadrants of the matrix. High-interest/high-influence stakeholders included the HSE, specialist centres, general practitioners, and the Endometriosis Association of Ireland. High-interest/low-influence groups comprised patients, families, and online communities, while policymakers, hospital managers, and the education sector were identified as high-influence but low-interest actors. Key drivers included strong patient advocacy, institutional support such as engagement from the HSE, and growing awareness of digital health tools. Major barriers encompassed prolonged diagnostic delays, resource constraints, gaps in clinical knowledge, technology anxiety, and challenges sustaining engagement.

**Conclusions:** Stakeholder mapping provided an evidence-informed foundation for the VendoR project, revealing engagement gaps and leverage points critical for improving endometriosis care innovation. The findings highlight the need for intentional, well-resourced strategies that elevate patient voices, address systemic barriers, and ensure balanced representation, supporting the co-design, co-creation, and co-production of digital health interventions for sustainable, patient-centred care.

## 1. Introduction

Endometriosis affects approximately 10% of reproductive-age women worldwide, leading to CPP(1) and significantly negatively impacts their quality of life (QoL) (2) and economic productivity. The EU estimates costs at €30 billion annually (3). Current treatments, primarily pharmacological and surgical, pose barriers such as high costs, accessibility issues (4), risks (5), complications (6), symptom recurrence, fertility impact (7), and lengthy recovery times.

Improving care pathways requires coordinated engagement of diverse stakeholders, including healthcare providers, policymakers, educators, technology developers, and patients themselves. A stakeholder is any individual, group, or organization that has an interest in, or may be affected by, a research study and its intended or unintended outcomes (8). It also encompasses those who may have an interest in the study or the power to influence its outcomes, either positively or adversely. A stakeholder does not have to be a direct user of the health intervention in development in order to be influenced or affected by the research or its outcomes. This means that a wide range of stakeholders can be involved in the co-design, co-creation, or co-production of research, such as: patients and caregivers, community members and leaders, healthcare providers, general public, policy makers, media, clinical leaders and associations. However traditional approaches to health innovation often fail to meaningfully and systematically involve stakeholders, limiting legitimacy, effectiveness, and uptake of interventions.

An increased number of studies demonstrate that stakeholder engagement enhances the relevance, quality, and overall impact of research and decision-making processes (9, 10). A systematic review of 66 studies demonstrated that patient and public involvement (PPI) enhances the quality and relevance of health and social care research across all stages, from shaping user-focused objectives and research questions to improving study materials, recruitment strategies, data interpretation, and the implementation and dissemination of findings, while also acknowledging some practical challenges (11). Participatory methods, including stakeholder mapping and deliberative workshops, are recognised as effective for capturing diverse perspectives, eliciting values, and identifying structural barriers in complex systems (12, 13). These approaches facilitate shared understanding, support co-creation, co-design and co-production of evidence, and inform decision-support frameworks in healthcare innovation (11, 13, 14).

The VendoR study aims to evaluate the usability and feasibility of a nature-based immersive Virtual Reality (<u>VR</u>) therapeutic intervention for pain management in women with <u>endo</u>metriosis. The project seeks to guide and inform the development of digital support tool for endometriosis care in Ireland by actively involving stakeholders as co-creators and co-producers throughout the research process. As co-creators, stakeholders will test the VR intervention and evaluate the surveys prior to pilot launch, providing feedback on usability, accessibility, and overall experience. Their insights will inform refinements to ensure the intervention is relevant and user-friendly for the target population. As co-producers, stakeholders will help disseminate study findings by acting as advocates and sharing their experiences with broader communities, supporting the communication of the intervention’s value and impact.

The project ran a stakeholder mapping workshop engaging participants from healthcare, policy, education, technology, and patient communities. The workshop aimed to identify the different stakeholders, assess their influence and interest, and to identify key drivers and barriers affecting the uptake of innovative approaches to endometriosis care. These insights are essential to designing inclusive, effective, and sustainable interventions that align with lived experience and systemic realities.

This paper presents the outcomes of this structured stakeholder mapping workshop undertaken to lay the collaborative foundations for project development.

## 2. Objectives

The workshop was designed to achieve the following objectives:

1. Identify and categorize key stakeholders relevant to endometriosis care in Ireland.
2. Assess each stakeholder’s level of influence and interest in the VendoR initiative as a digital support tool for pain management.
3. Map stakeholders on an Influence vs. Interest Matrix to visually represent engagement dynamics.
4. Identify drivers and barriers affecting the uptake of innovative approaches to endometriosis care.

## 3. Methods

### 3.1 Workshop Design

A virtual stakeholder mapping workshop was conducted on 17 June 2025, facilitated by members of the VendoR research team at SETU,, Walton Institute in collaboration with SETU Research, Innovation and Impact (RII) office. The session aimed to gather perspectives of key stakeholders, their roles, and explore drivers and barriers influencing the uptake of endometriosis care innovations.

Two participatory tools were used:

- **Stakeholder Mapping Matrix:** Stakeholder Mapping Matrix: In a collaborative setting, participants first identified all relevant stakeholders in endometriosis care across categories such as the healthcare system, policymakers, the educational sector, and endometriosis support communities, not-for-profit-volunteer-run organisation (Figure 1: *All the names provided during this workshop are covered for privacy*). Participants then collaboratively positioned these stakeholders on a matrix according to their levels of influence and interest (15) (Figure 2).
- **Drivers and Barriers Board:** Participants contributed qualitative reflections via virtual sticky notes, highlighting factors enabling or constraining uptake of endometriosis care innovation. The workshop was delivered in a structured format using MS Teams whiteboard, with breakout discussions and plenary reflection.

**Figure 1.**
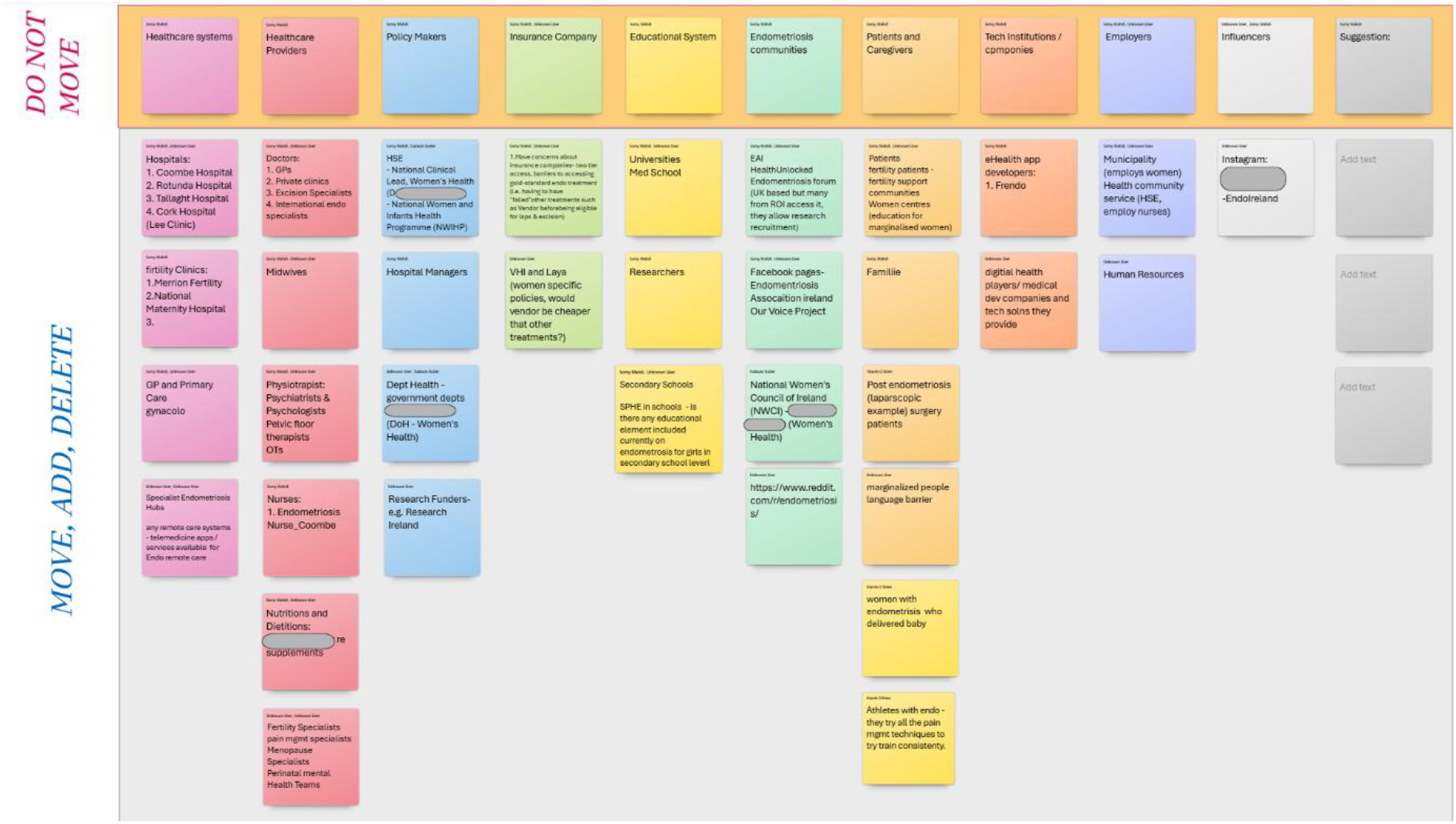
All stakeholders related to endometriosis care.

**Figure 2:**
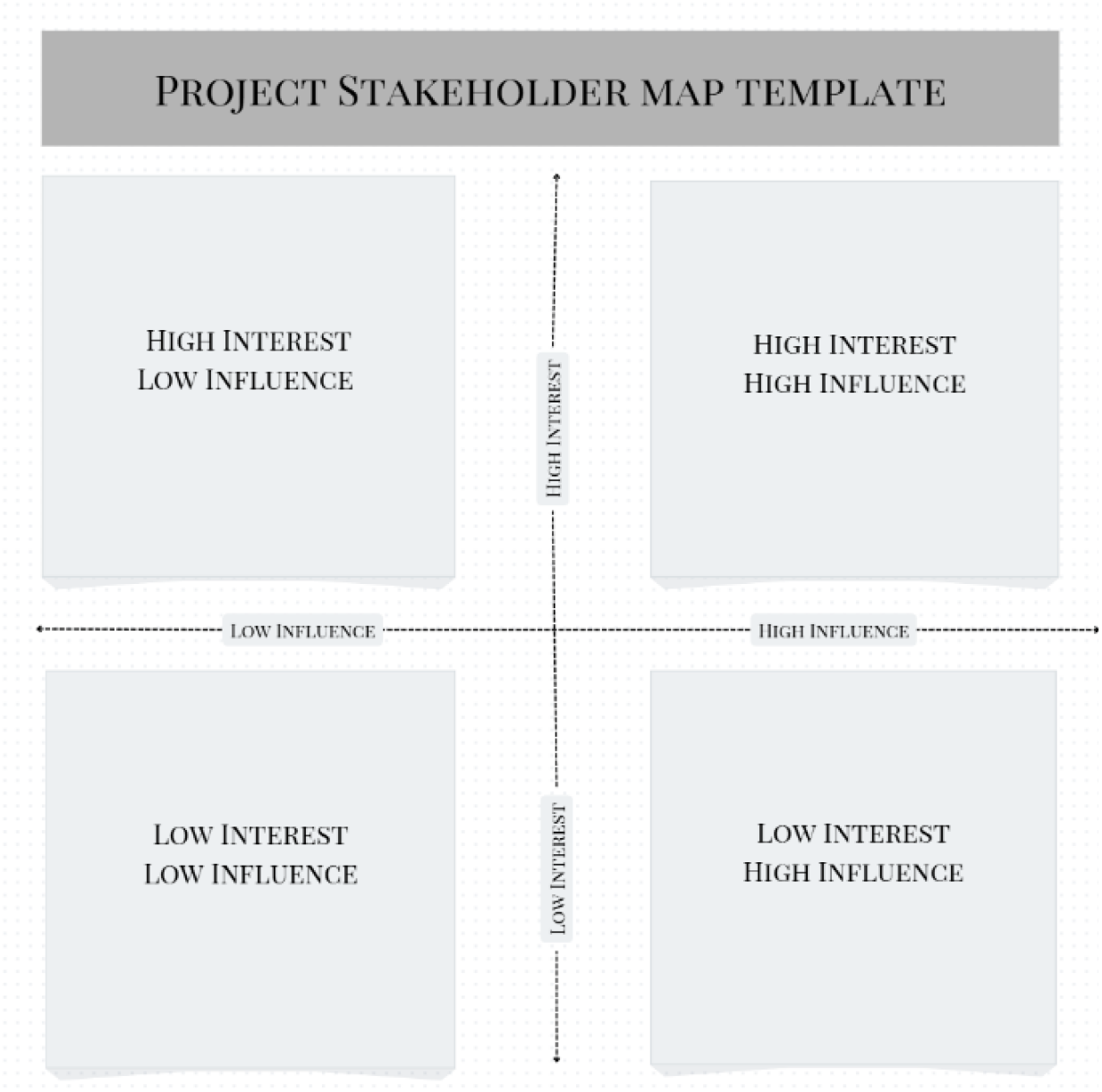
Stakeholder Mapping Template.

### 3.2 Participation and Composition

Participation was open and included a broad range of participants, including individuals with lived experience, clinicians, researchers, students, digital health developers, and professionals from mental health and allied health sectors. The workshop detail and an open invitation to participate was circulated via social media, institutional communications, and a dedicated project webpage. This ensured diverse perspectives reflecting the realities of endometriosis care.

### 3.3 Ethical Considerations

The workshop was non-clinical and non-interventional. Key ethical measures included voluntary participation, anonymized contributions, no audio or video recording, and reporting of only aggregated findings. Although the activity was conducted outside the scope of formal ethics review, it adhered to standard participatory research guidelines. The VendoR Project has received full ethical approval from the SETU Ethics Committee (reference numbers: SETU/REC/25/26/012 and SETU/REC/25/26/046).

## 4. Results

### 4.1 Stakeholder Categories and Matrix Placement

Stakeholders were distributed across four quadrants of the influence–interest matrix:

- **High Interest / High Influence:** Health Service Executive (HSE), specialist centres, general practitioners, midwifery services, national advocacy leaders, the Endometriosis Association of Ireland (EAI), primary care gynaecology, hospitals and women’s health centres, and the National Women and Infants Health Programme (NWIHP).
- **High Interest / Low Influence:** Patients, families, online support communities, universities and researchers, social media groups, and athletes living with endometriosis.
- **Low Interest / High Influence:** Hospital managers, policymakers, insurers, the education system (including secondary schools and Social Personal Health Education (SPHE)), healthcare providers such as nurses and specialist endometriosis physiotherapists, and the Department of Health.
- **Low Interest / Low Influence:** HR departments, dietitians, and other peripheral or disconnected actors.

**Table 1.**
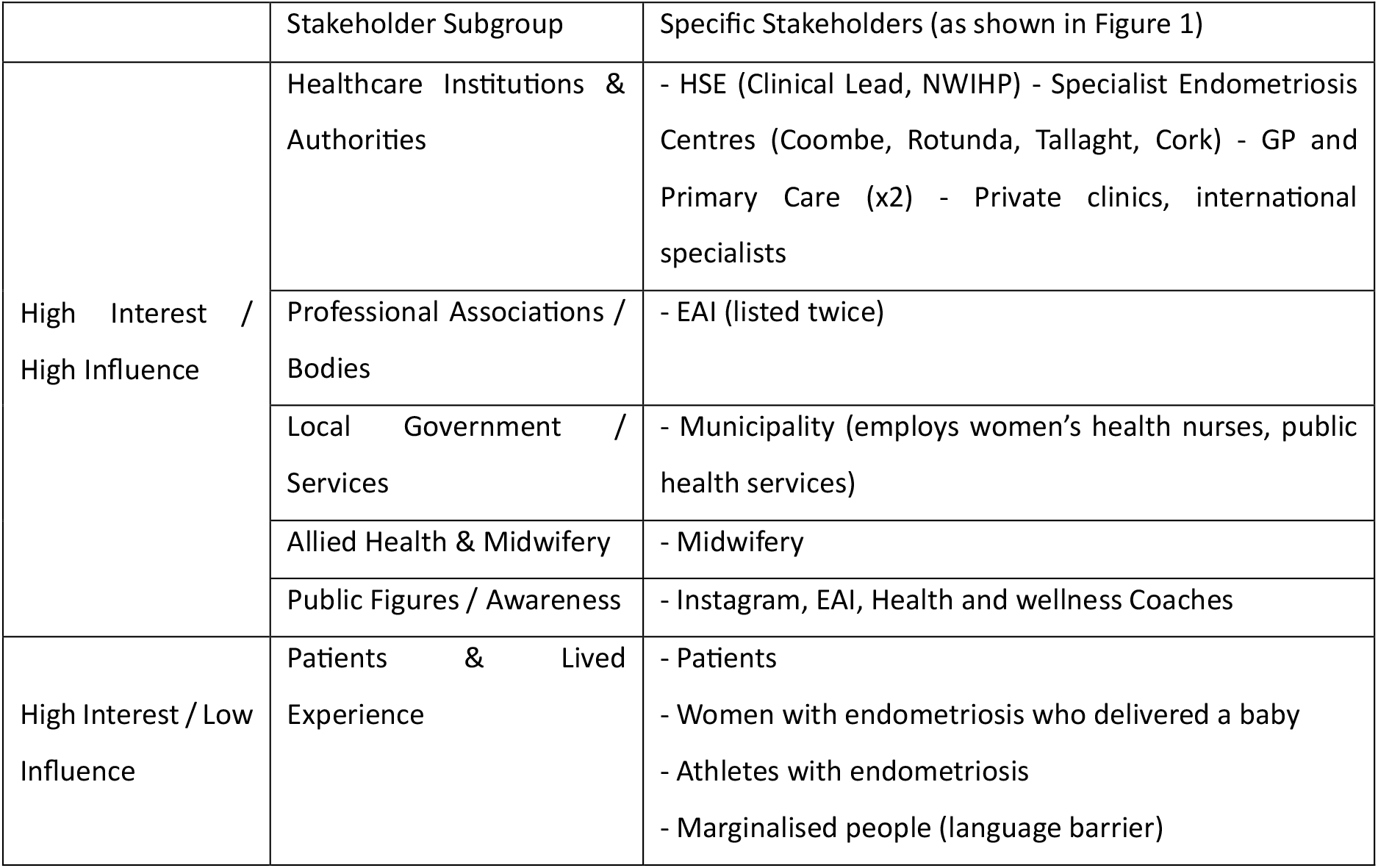
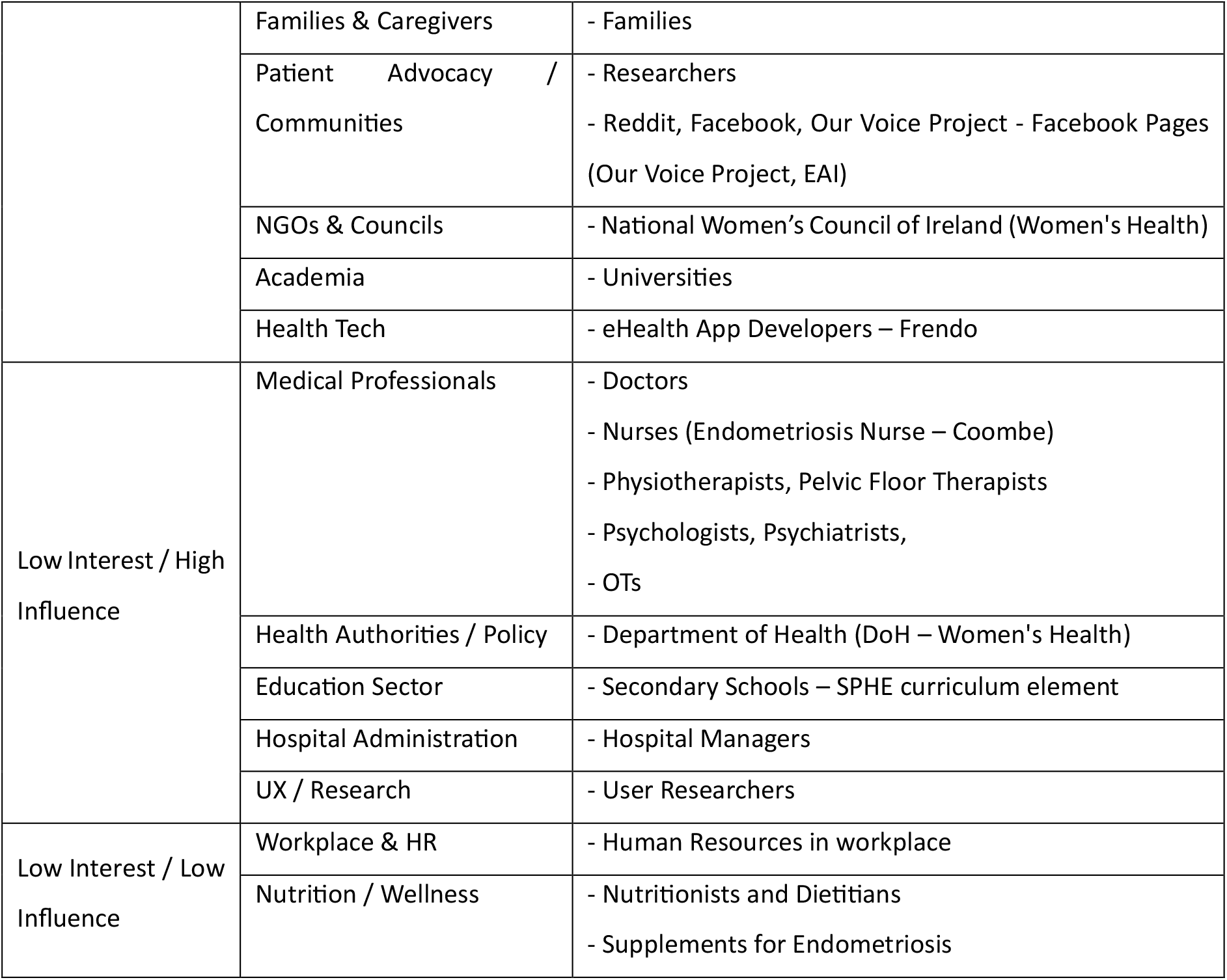
summarises all stakeholder categories and their final placement in the matrix.

### 4.2 Drivers and Barriers to Stakeholder Engagement

#### Drivers

Key drivers for the uptake of innovative approaches to endometriosis care included strong patient advocacy and motivation, with individuals with lived experience and patient organisations willing to engage with well-communicated, evidence-based interventions that build trust and empowerment. Institutional and systemic support, including public funding, engagement from the HSE, and the potential to demonstrate public health value through measures such as Social Return on Investment, were also identified as important enablers.

Access to centralised, verified information was viewed as essential for supporting self-management and countering misinformation. In addition, the involvement of primary care professionals, particularly GPs and frontline healthcare providers, was recognised as critical for early identification, management, and referral, and therefore for successful uptake of innovation.

Drivers of mild importance included increasing awareness of digital health technologies and innovation, supported by growing familiarity with emerging tools and platforms already available in market, as well as greater visibility of technology-enabled care through healthcare conferences and research dissemination. Allied and community-based health services, including women’s physiotherapy and specialised clinics such as postnatal care programmes, were identified as valuable yet underutilised contributors to awareness, recovery, and continuity of care. In addition, participants highlighted the importance of improved information access and community voice, expressing a desire for centralised platforms to share feedback and lived experiences. Carers and patients emphasised the need for meaningful involvement, with existing peer networks and social media platforms recognised as channels that could be more effectively leveraged for engagement and dissemination.

Drivers of low importance included the growing recognition of the need to elevate the patient voice and to challenge systemic gender bias in healthcare. Participants acknowledged increasing attention to patient narratives in public discourse, citing recent advances in menopause awareness as a potential reference point for endometriosis advocacy. However, while these cultural shifts were viewed as important for long-term transformation, they were considered less immediately influential on the short-term uptake of innovative endometriosis care solutions. Similarly, entrenched gender bias was recognised as a structural issue contributing to underdiagnosis, symptom dismissal, and limited policy prioritisation, yet stakeholders perceived meaningful change in this area as gradual and beyond the direct control of individual innovation initiatives.

#### Barriers

Barriers of high importance were primarily systemic and structural, significantly limiting the uptake of innovative approaches to endometriosis care. Prolonged diagnostic delays, often spanning 7-20^+^ years, along with extensive waiting lists and slow ethics, governance, and approval processes for innovation care research, were identified as major obstacles to timely innovation adoption. Resource and budget constraints, including limited protected time for clinicians, restricted public healthcare funding, and reliance on costly private services, further exacerbated inequalities in access and constrained participation in innovation initiatives. Participants also highlighted substantial gaps in clinical knowledge of endometriosis, particularly within primary care, which hinder early identification and reduced confidence in adopting new models of care or digital interventions. In addition, the cognitive and physical burden associated with endometriosis, such as chronic pain, fatigue, brain fog, and emotional exhaustion, was recognised as a critical barrier to patients’ sustained engagement with innovative tools, co-production activities, and decision-making processes.

Barriers of mild importance related to accessibility, engagement, and implementation processes. Digital and physical accessibility challenges, including technology anxiety, limited access to appropriate devices, lack of private space at home, and scalability concerns, were identified as barriers to equitable uptake, particularly among socioeconomically disadvantaged groups. Gaps in informal and community support systems, including limited understanding among family members and low public awareness of endometriosis, were seen to reduce acceptance and sustained use of innovative care approaches. Participants also reported research and consultation fatigue, noting repeated requests for input without visible impact or feedback, which undermines trust and willingness to engage with innovation initiatives. Concerns around data governance and consent processes, particularly in relation to GDPR compliance, patient data access, and the complexity of consent and re-consent mechanisms, were identified as additional barriers to engagement with digital health innovations.

Barriers of low importance reflected cultural and organisational challenges affecting long-term innovation uptake. Resistance to change among some healthcare professionals and institutions was noted, particularly where existing practices were perceived as adequate, alongside fragmentation across primary care, hospital services, specialist centres, and community organisations. Participants also highlighted a disconnect between research priorities and lived experience, where innovation agendas may overlook patient-defined needs, leading to limited relevance and adoption. Finally, frustration was expressed regarding repeated acknowledgement of systemic issues without tangible implementation, reinforcing scepticism toward new initiatives and highlighting the need for action-oriented, patient-informed innovation pathways.

## 5. Discussion

The VendoR stakeholder mapping workshop highlighted the critical importance of aligning positional power with lived experience in endometriosis care innovation. The identification of highly interested but low-influence stakeholders, such as patients, caregivers, and online support communities, underscores the need for strategies that redistribute influence through co-design, inclusive communication, and empowerment of underrepresented voices. This aligns with previous work demonstrating that structured stakeholder engagement improves relevance, legitimacy, and uptake of research projects by ensuring that decision-making incorporates multiple perspectives and lived experience (10, 16, 17).

Visual and thematic mapping of stakeholders also provided insights into systemic gaps in endometriosis care in Ireland. High-influence but low-interest stakeholders, such as policymakers, hospital administrators, and insurers, represent critical targets for engagement to ensure alignment of resources, policy, and organizational support. This observation is consistent with participatory systems approaches in health research, which stress the need of engaging actors across varying levels of influence and interest to identify leverage points and potential bottlenecks in care pathways (18, 19).

The workshop also highlighted barriers and facilitators relevant to stakeholders engagement. Technology Anxiety although ranked as a mild barrier in the workshop, ‘tech anxiety’ emerged in academic literature as a significant obstacle to digital health adoption, especially among older adults and patients with chronic conditions. Studies show that technology anxiety negatively influences ease-of-use and perceived usefulness, reducing behavioural intention to engage with digital health tools (20). Addressing this barrier is essential, particularly in projects like VendoR that rely heavily on digital tools and remote care modalities.

Drivers such as strong patient advocacy, institutional support from the Health Service Executive, access to verified information, and involvement of primary care providers were identified as high-impact enablers, which mirrors findings in broader evidence synthesis research emphasizing that early and sustained stakeholder engagement enhances the quality, relevance, and implementation of health interventions (16, 21).

Overall, these findings demonstrate that stakeholder mapping provides a structured, evidence-informed approach to identify key actors, potential engagement gaps, and systemic barriers. By combining visual mapping with qualitative insights on drivers and barriers, the workshop delivered actionable intelligence to inform the next stages of the VendoR project, particularly co-design of digital tools, policy engagement, and education strategies.

While stakeholder engagement is essential for developing relevant, patient-centred innovations in endometriosis care, this process also presents practical and methodological challenges that must be acknowledged in future phases of the VendoR project. Effective engagement requires time, resources, and deliberate planning to avoid tokenistic participation and ensure balanced representation across diverse groups. Clear communication of objectives, structured processes for managing conflicting perspectives, and adequate support for both stakeholders and researchers are critical to sustaining meaningful involvement.

Confidentiality, equitable attribution of contributions, and safeguards against undue influence must remain central considerations. By anticipating and addressing these challenges, the VendoR project is well positioned to build a robust, inclusive co-creation and co-production process that strengthens the legitimacy, feasibility, and long-term impact of digital health innovation in endometriosis care.

## 6. Conclusion

This study demonstrates that structured stakeholder mapping is a valuable tool for guiding innovation in endometriosis care. By systematically identifying and categorizing stakeholders, assessing their influence and interest, and capturing drivers and barriers affecting the uptake of innovative approaches to endometriosis care, the VendoR project was able to uncover critical gaps and leverage points within the care ecosystem.

High-interest but low-influence stakeholders, such as patients and community groups, require intentional strategies to amplify their voices, while high-influence but low-interest actors, including policymakers and hospital administrators, represent key targets for engagement to ensure alignment of resources and policy support. The findings underscore the importance of early, well-resourced, and inclusive engagement processes that integrate lived experience, professional expertise, and systemic insights.

Anticipating structural barriers, resource limitations, and digital adoption challenges is essential for sustaining meaningful participation and ensuring that innovations are feasible, relevant, and impactful. Overall, this work provides an evidence-informed foundation for co-designing, co-creation, or co-production digital health interventions, informing policy and educational strategies, and advancing patient-centred endometriosis care in Ireland.

Future research should continue to explore mechanisms for balancing influence and interest among stakeholders, addressing systemic barriers, and evaluating the long-term impact of participatory approaches on the uptake and sustainability of innovative health solutions.

## Data Availability

All data produced in the present work are contained in the manuscript

## Notes

### Competing Interest Statement

The authors have declared no competing interest.

### Funding Statement

This study was funded by SET TU RISE

### Author Declarations

The VendoR Project has received full ethical approval from the SETU Ethics Committee (reference numbers: SETU/REC/25/26/012 and SETU/REC/25/26/046).

